# Determinants of enrollment into community based health insurance among households in Chora District, Southwest Ethiopia: Unmatched Case Control Study

**DOI:** 10.1101/2023.12.30.23300668

**Authors:** Awolu Abdulkadir Disasa, Tadesse Gebremedhin, Mohammad Jihad

## Abstract

**Background:** Community-based health insurance (CBHI) has emerged with exciting ideas in accessing reliable health care services for low and middle-income countries. Despite increasing effort, empirical evidence shows enrolment has remained low in Ethiopia. Therefore, this study aimed identifies determinants of enrollment into CBHI in Southwest Ethiopia.

**Methods:** Community based unmatched case-control study was conducted in Chora district, Bunno Beddele Zone Southwest Ethiopia from 17th May-14th June 2021. About 245 cases and 245 controls (1: 1 ratio) were selected by simple random sampling technique, and participated in the study. Data were collected using face-to-face interview techniques with pre-tested questionnaire, and entered to Epi-data 4.6, then exported to SPSS version 25 for analysis. Logistic regression analyses were conducted to identify risk factors associated with enrolment into CBHI. The strength of association was measured using odds ratio. The findings were presented along with their corresponding 95%CI. Variables with p-values <0.05 were considered as determinants of CBHI enrolment.

**Results:** The study revealed that female-headed households [AOR:10.86, 95%CI: 3.005-39.16], age of household head, being 35-50 years old [AOR:3.81, 95%CI:1.61-9.02], and being above50 years [AOR: 3.59, 95%CI: 1.23-10.47], having 4-5 family sizes [AOR:7.3, 95%CI: 2.08-25.84] and ≥6 family sizes [AOR:13.31, 95%CI: 3.86-45.92], distance from health facility of ≤30 minutes [AOR:8.35, 95%CI: 3.22-21.65], favorable perception toward providers technical competencies [AOR:7.62, 95%CI:2.2-26.4], fully trusted on CBHI scheme [AOR:10.82, 95%CI:2.37-45.17] and moderate trust towards CBHI scheme managements [AOR:4.91, 95%CI:1.9-12.65] and positive attitudes towards CBHI scheme [AOR:4.34, 95%CI: 1.525, 12.32] were determinants among households for CBHI enrollment.

**Conclusion:** Availability of contractual health-facility at nearest distances, being households’ head aged above 35years, being female-headship, large family-size, favorable perception towards health care providers technical competencies, trust towards scheme management, and positive attitudes toward scheme were determined CBHI scheme enrollment.

## Introduction

Community-based health insurance (CBHI) is a pledge agreement requiring the health insurer to cover basic health service costs in exchange for premium payments into a collective fund which is designed, owned, and administered by members (1). It was a complementary source of health care finance that has been implemented as part of health reform programs and aimed at improving the health status of the poor by protecting them from the financial hardship through risk pooling across members (2–6). It improves health service utilization, and help to extend coverage to rural and the informal sector workers through providing financial protection among members by reducing their out-of-pocket payments (OOPPs), and helps to contribute for universal health coverage (7–9).

Globally the resource-poor settings such as low and middle income countries (LMICs) have existed with very weak health care systems, and poor health-seeking behaviors with a high burden of diseases due to inaccessibility and poor utilization of health care facilities accompanied by different factors. Study by World Health Organization (WHO) stated that due to out-of-pocket payments (OOPPs) on health services every year, about 100 million people have been pushed into poverty, and 150 million people suffer financial catastrophe (10).

In developing countries over 2 billion people are living with health systems afflicted by inefficient, inequitable access, inadequate funding and poor quality of health care services (11,12). However, financial barriers were being the major bottleneck for not accessing and non-using health services, and health-related financial stocks are further exacerbated when the primary income earner in poor households are the ones that require the expensive health care treatments (13).

In most developing countries, still health care cost is mainly paid at a time of sickness and OOPPs at the point of service delivery that potentially inhibiting accessibility. However, increasing coverage with voluntary health insurance in low-income settings is challenging due to low CBHI uptake among households of informal sector workers (14,15). In such a countries, initiatives are underway to strengthen voluntary CBHI as a means of expanding access to affordable care among the informal sector and as an emerging tool for providing financial protection against health-related poverty (14–16). In Ethiopia, the national CBHIs memberships among households were only 49% at the end of Health Sector Transformation Plan (HSTP) I by 2020 (17). African populations are still relying mostly on OOPPs(accounting for 30%-85% of total health spending) and are associated with a higher probability of incurring very expensive health expenditure and impoverishment (11). Similarly, in Ethiopia, OOPPs was 37% in 2015/2016 and was planned to reduce to < 25% by 2020 (18).

In Ethiopia, despite high burden of infectious and non-infectious diseases, the modern health care services utilization of were limited due to user-fee charges (25). Therefore, to avert these difficulties, CBHI scheme was launched as the solution, and its enrollment rate was found to be promising as compared to other African countries (16,26). The Ethiopian health insurance agency (EHIA) evaluation finding showed that the enrollment rate into CBHI has been 44% in 509 CBHI functional districts (16). There are also regional variations in enrollment, and contribution of annual premium until 2019 (25%) at Benshangul-Gumuz region, and in Oromiya region (33%) (16). On the other hands, a few studies were also conducted on factors associated with enrollment into CBHIs scheme (10,21,27,28).

Moreover, even though there are few studies conducted in the country, most of them were cross-sectional studies mainly focusing on willingness to join (WTJ) CBHI scheme. Few studies that were conducted on determinants of enrollment into CBHI were mostly confined to the Northern parts of the country; which is not similar to the current study area in different aspects like cultural and socio-demographic situations. Therefore, the current study was aimed to determine factors that are responsible for CBHI enrolments among households in Chora district Southwest Ethiopia.

## Methods

### Study design and setting

A community-based unmatched case-control study design was conducted from 17^th^ May-14^th^ June 2021in Chora district, Bunno Beddele Zone, Oromia Regional State, Southwest Ethiopia. CBHI activities were officially launched in Nov 2018 in all Kebeles, of the zone. Between 2018 and 2020, only 55% of the households were enrolled into CBHI scheme.

### Source and study populations

The source and study population for the cases were all households who were enrolled in to community-based health insurance in the study area (self-paid and subsidized households) while households who were not enrolled into CBHI schemes in the study are were the source and study populations for the controls.

### Inclusion criteria and Exclusion criteria

For cases: households heads who are registered at villages (called Kebele) level on membership registration logbooks and enrolled into CBHI scheme, renewed their membership and households who have paid annual premium.

For controls: Household heads that were not enrolled into CBHI scheme in the study area and lived in the study area for at least 6 months prior to data collections were included. Household heads that were seriously ill and/or unable to communicate during data collection were excluded from the study.

### Sample size

The required sample size was determined using Epi-info version 7 based considering parameters of two population proportion formulas: 5% precision, 95% confidence interval, 80% power and 1: 1 ratio of controls to cases, Moreover, by assuming 7.4% of merchant household heads will not be enrolled into CBHI scheme (controls) in the study area (28) and non-response rate of 5%, the 245 cases and 245 controls household heads were recruited into the study.

### Sampling procedure

We selected 30% of the Kebeles (12 Kebeles) using simple random sampling technique. Then, the sample size was proportionally allocated to each selected Kebele based on their existing number of cases and controls in each Kebele. We prepared a sampling frame of cases and controls using Community Health Information System (CHIS) found at each Kebele. Then, we applied computer generated ransom number (generated using excel command) to select the cases and controls.

### Data collection procedures

Structured, pretested questionnaire was to collect data form participants. Data were collected by persons who completed 10^th^ grade and supervised by public health professionals. Supervisors and principal investigator reviewed and checked data for errors (outliers, inaccuracies, missing etc.).

### Data processing and analysis

Data were edited and coded; then entered into Epi data version 4.5. Data were cleaned and analyzed using SPSS version 25. We computed descriptive statistics and presented them using frequency tables, and percentages. Bivariable logistic regression analyses were performed to identify candidate variables at *p*-value ≤0.25. Multivariable logistic regression analysis were also performed to identify factors associated with the outcome variable at *p*-value <0.05. Finally, the findings were presented using crude and adjusted odds ratios along with corresponding 95%CIs goodness of fit of the model was assessed using Hosmer and Lemeshow statistics; and overall prediction power of the model 93.3%.

### Ethical consideration

Ethical approval was received from institutional ethical review board of Jimma University, Institute of Health science. Chora district gave us the permission letter for every 12 Kebeles. Kebeles confirmed our letter of permission and then the data collector took permission to collect data from each household. After explaining the overall objectives of the study, verbal informed consent was received form each participant. The respondents’ right to refuse or withdraw from participating at any time was fully respected and the information provided by each respondent was kept confidential by making each questionnaire coded and not sharing personal information of any patient to the third party.

## Results

### Socio-demographic characteristics of house-hold head

All household heads participated in the study (100%). One hundred ninety four (79.2%) of cases and 226 (92.8%) of controls were male (Table 1). The mean of the age of cases and controls were 44.06 (±1.55 SD) and 36.33(±1.57 SD) years respectively. However, among controls the mean ages were years. About 203 (82.9%) among cases and 146 (59.6%) among controls were married. On other hands, 101 (41.2%) of cases and 164 (66.9%) of controls were unable to read and write.

**Table 2:**
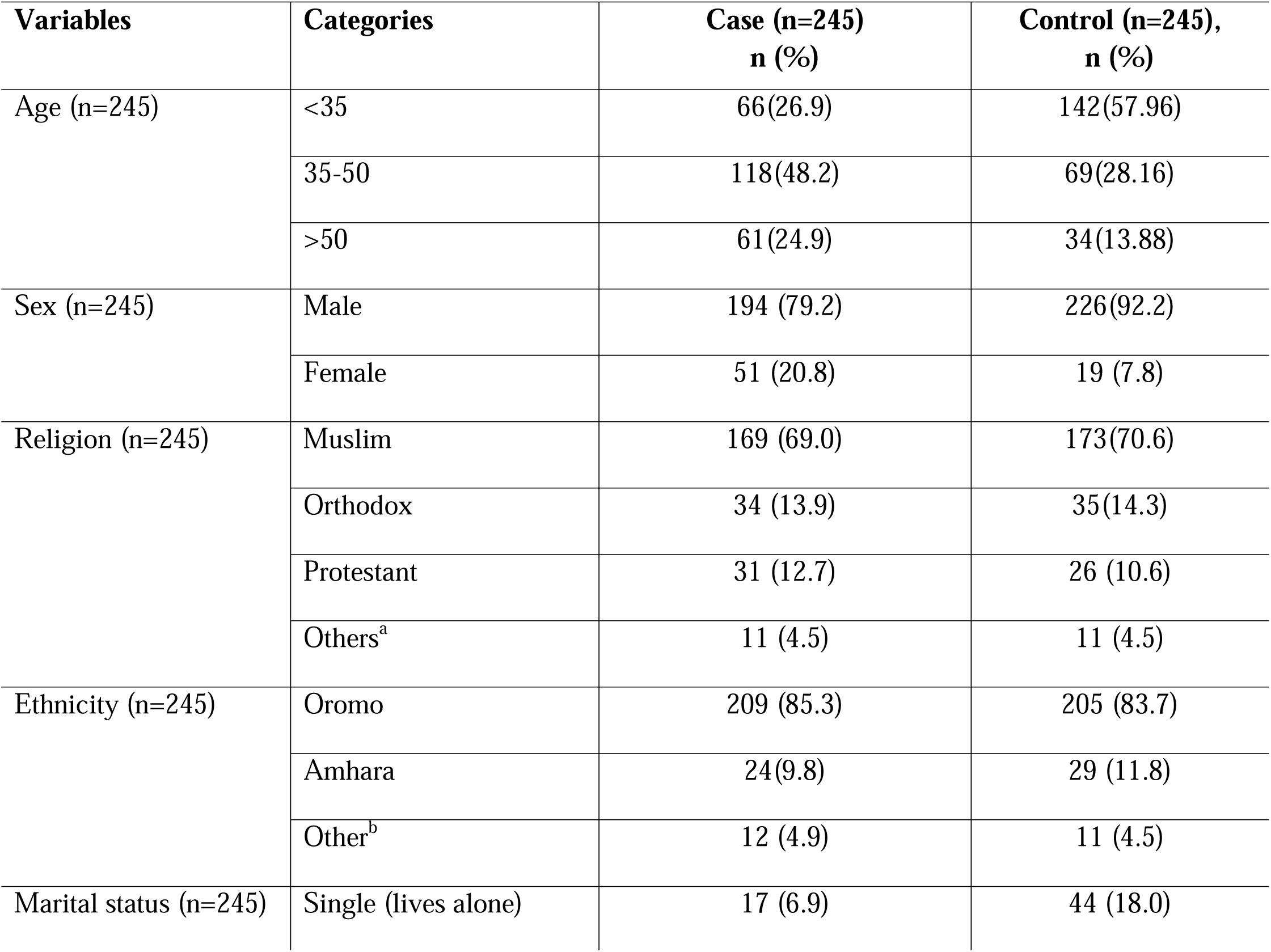
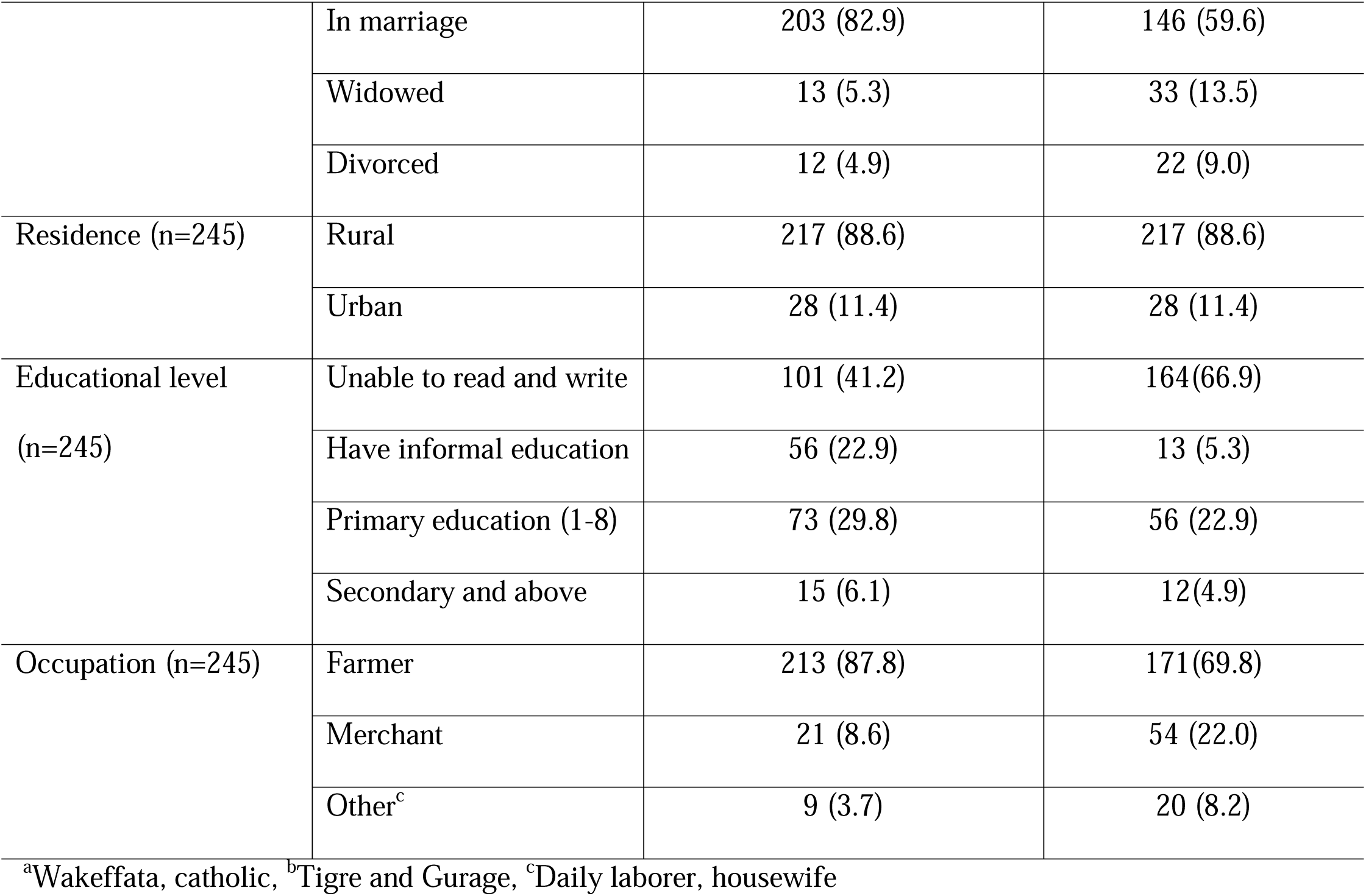
Socio-demographic characteristics among households in Chora District, Southwest Ethiopia, 2021.

### Household level related characteristics

About 151 (61.6%) of cases and 49 (20.0%) of controls were had ≥6 family size while 15 (6.1%) of cases and 101 (41.2%) of controls had less <3 family sizes (Table 3). About 45(18.4%) cases and 6.1% (15) of controls respectively had encountered medically proved chronic illnesses. About 8.6% (21) of cases and 38.4% (94) of controls had perceived to have a very good family health status. More than half among cases 137(55.9%) and controls 139(56.7%) were found in the 4^th^ wealth Quintiles.

**Table 4:**
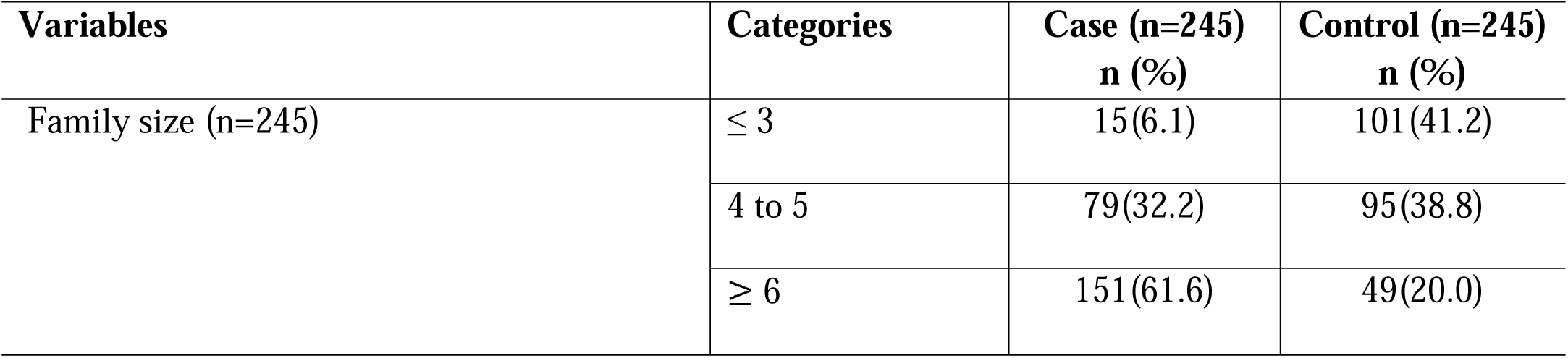
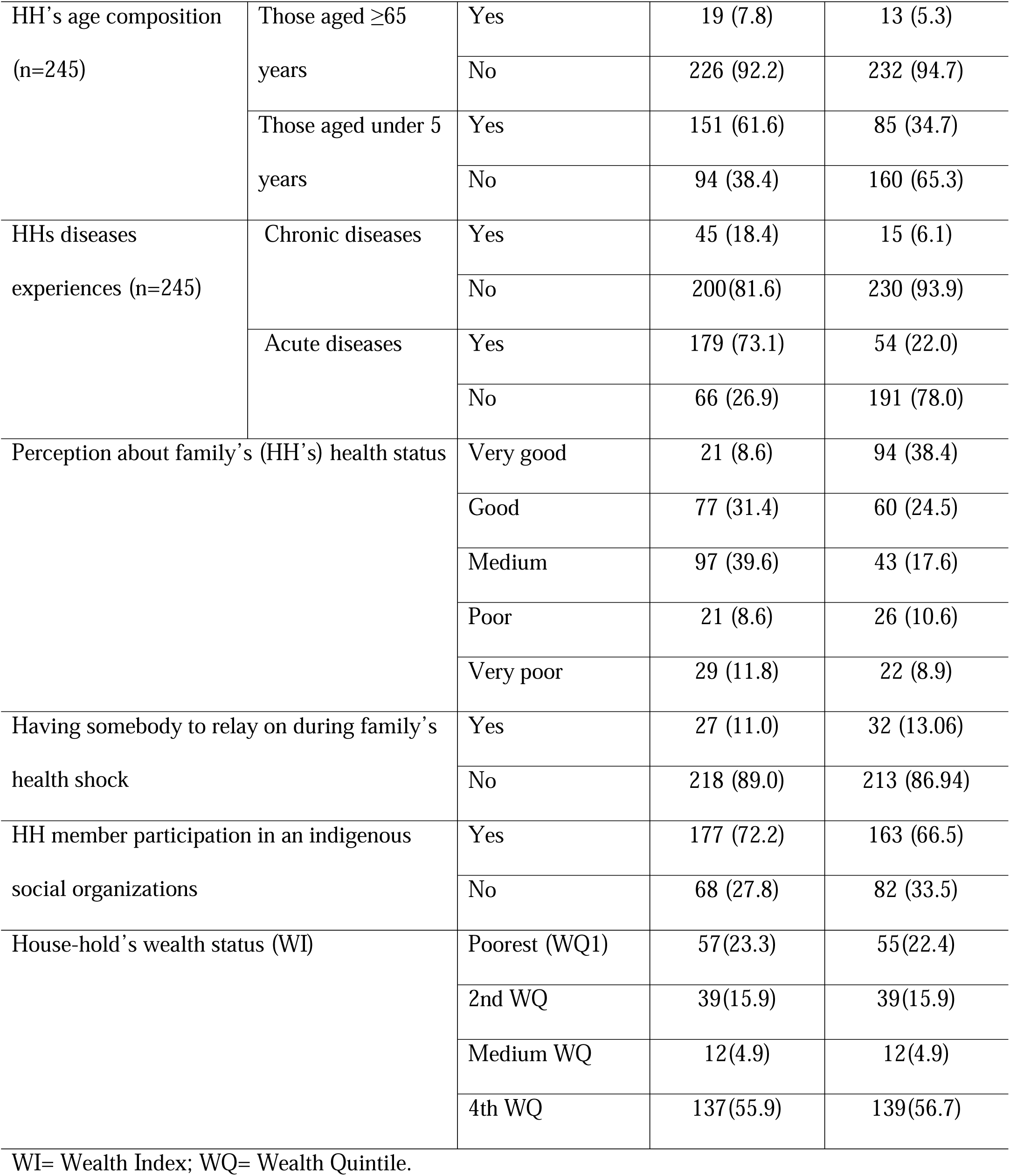
Chora District, Southwest Ethiopia, 2021.

| Variables | Categories | Case (n=245)<br>n (%) | Control (n=245)<br>n (%) |
| --- | --- | --- | --- |
| Family size (n=245) | $\leq 3$ | 15(6.1) | 101(41.2) |
|  | 4 to 5 | 79(32.2) | 95(38.8) |
| | $\geq 6$ | 151(61.6) | 49(20.0) |

|  |  |  |  |  |
| --- | --- | --- | --- | --- |
| HH's age composition<br>(n=245) | Those aged $\geq 65$<br>years | Yes | 19 (7.8) | 13 (5.3) |
|  |  | No | 226 (92.2) | 232 (94.7) |
|  | Those aged under 5<br>years | Yes | 151 (61.6) | 85 (34.7) |
|  |  | No | 94 (38.4) | 160 (65.3) |
| HHs diseases<br>experiences (n=245) | Chronic diseases | Yes | 45 (18.4) | 15 (6.1) |
|  |  | No | 200(81.6) | 230 (93.9) |
|  | Acute diseases | Yes | 179 (73.1) | 54 (22.0) |
|  |  | No | 66 (26.9) | 191 (78.0) |
| Perception about family's (HH's) health status | Very good |  | 21 (8.6) | 94 (38.4) |
|  | Good |  | 77 (31.4) | 60 (24.5) |
|  | Medium |  | 97 (39.6) | 43 (17.6) |
|  | Poor |  | 21 (8.6) | 26 (10.6) |
|  | Very poor |  | 29 (11.8) | 22 (8.9) |
| Having somebody to relay on during family's<br>health shock | Yes |  | 27 (11.0) | 32 (13.06) |
|  | No |  | 218 (89.0) | 213 (86.94) |
| HH member participation in an indigenous<br>social organizations | Yes |  | 177 (72.2) | 163 (66.5) |
|  | No |  | 68 (27.8) | 82 (33.5) |
| House-hold's wealth status (WI) | Poorest (WQ1) |  | 57(23.3) | 55(22.4) |
|  | 2nd WQ |  | 39(15.9) | 39(15.9) |
|  | Medium WQ |  | 12(4.9) | 12(4.9) |
|  | 4th WQ |  | 137(55.9) | 139(56.7) |

WI= Wealth Index; WQ= Wealth Quintile.

### Health services related characteristics among cases and controls

About 215(87.8%) among cases and 218(89.0%) among controls got health center at their nearest to access, the nearest distances from their residences (Table 5). However about 97(39.6%) of cases and 38(15.5%) of controls reach health facilities with in ≤ 30 minutes a distance. The proportion of having good perceptions toward public facilities provider’s technical competencies were 96(39.3%) among cases and 45(18.4%) among controls. On the other hands, the proportions for having high satisfactions to perceived health care facilities’ service qualities were only 16(6.5%) among cases and 14(5.7%) among controls.

**Table 6:** Health care facility related characteristics for enrollment to CBHI among households in Chora District, Southwest Ethiopia, 2021.

| Variables | Categories | Case (n=245)<br>n (%) | Control (n=245)<br>n (%) |
| --- | --- | --- | --- |
| The nearest HF to your home that is usually used for medical care (n=245) | Health center | 215 (87.8) | 218 (89.0) |
|  | Hospitals | 16 (6.5) | 13 (5.3) |
|  | Other | 14 (5.7) | 14 (5.7) |
| Transportation cost from home to your HF (n=245) | Costly | 135 (55.1) | 125 (51.0) |
|  | Medium | 38 (15.5) | 36 (14.7) |
|  | Cheap | 72 (29.4) | 84 (34.3) |
| Distance to reach nearest HCFs (n=245) | $\leq 30$Mint' | 97(39.6%) | 38(15.5%) |
| | $>30$Mint' | 148(60.4%) | 207(84.5%) |
| Perceived health providers technical competences (n=245) | Favorable | 96 (39.2) | 45 (18.4) |
|  | Moderate | 133(54.3) | 126 (51.4) |
|  | Un-favorable | 16 (6.5) | 74(30.2) |
| Providers and client interactions (n=245) | Good | 59 (24.1) | 43 (17.6) |
|  | Neutral | 161 (65.7) | 157 (64.1) |
|  | Poor | 25 (10.2) | 45 (18.4) |
| Features of public health care facilities (n=245) | Good features | 37 (15.1) | 28 (11.4) |
|  | Neutral | 176 (71.8) | 169 (69.0) |
|  | Poor features | 32 (13.1) | 48 (19.6) |
| Satisfactions to health care facilities/ service qualities (n=245) | Highly satisfied | 16 (6.5) | 14 (5.7) |
|  | Moderate | 178 (72.7) | 137 (55.9) |
|  | Un-satisfied | 51 (20.8) | 94 (38.4) |

### Community based health insurance system related characteristics among cases and controls

The majorities among cases 213(86.9%) and among controls 182(74.3%) were found to have awareness about the scheme (Table 7). But about 78(31.8%) of cases and 23(9.4%) of controls were perceived the CBHI package to be adequate to meet their health care needs, and the largest proportions were in the middle to decide about its adequacy. However, about 3/4^th^ among both groups believed as CBHI scheme membership contributions premium was affordable to them. The proportion of having good knowledge about CBHI package benefits was about 181(73.9%) among cases and about 88(35.9%) among control. The proportions of having positive attitudes towards the scheme were about 176(71.8%) among cases and 42(17.2%) among controls.

**Table 8:** Scheme related characteristics for enrollment to CBHI among households in Chora District, Southwest Ethiopia, 2021.

| **Variables** | **Categories** | **Case (n=245)**<br>**n(%)** | **Control (n=245)**<br>**n(%)** |
| --- | --- | --- | --- |
| Awareness on CBHI (n=245) | Yes | 213 (86.9) | 182 (74.3) |
|  | No | 32 (13.1) | 63 (25.7) |
| CBHI package adequacy (n=245) | Adequate | 78 (31.8) | 23 (9.4) |
|  | Medium | 137 (55.9) | 119 (48.6) |
|  | Not Adequate | 30 (12.2) | 103 (42.0) |
| Trust to CBHI Scheme (n=245) | Trusted | 84 (34.3) | 25 (10.2) |
|  | Neutral | 134 (54.7) | 117 (47.8) |
|  | Not trusted | 27 (11.0) | 103 (42.0) |
| CBHI membership premium contribution<br>time interval (n=245) | Appropriate | 151 (61.6) | 146 (59.6) |
|  | Neutral | 65 (26.5) | 61 (24.9) |
|  | Not appropriate | 29 (11.8) | 38 (15.5) |
| Affordability of membership<br>contributions (n=245) | Affordable | 195 (79.6) | 183 (74.7) |
|  | Neutral | 29 (11.8) | 31 (12.7) |
|  | Not affordable | 21 (8.6) | 31 (12.7) |
| Knowledge toward CBHI (n=245) | Poor | 11(4.5) | 57(23.3) |
|  | Moderate | 53(21.6) | 100(40.8) |
|  | Good | 181(73.9) | 88(35.9) |
| Attitudes toward CBHI (n=245) | Positive | 176 (71.8) | 42 (17.2) |
|  | Neutral | 35 (14.3) | 91 (37.1) |
|  | Negative | 34 (13.9) | 112 (45.7) |

### Determinant factors for CBHI enrollment decisions among households in Chora district

In multivariate logistic regression analysis, family size, age, distance to reach health facility, perceptions toward provider’s technical competencies, trust towards CBHI scheme management and attitude towards CBHI scheme were the risk factors that associated with households enrollment into CBHI scheme (p<0.05). The study depicted that the odds of enrolment into CBHI scheme among female-headed household were 10.86 times higher as compared to male-headed households [AOR: 10.86, 95%CI: 3.01 - 39.16] (Table 9). The odds of enrolment into CBHI scheme among household heads aged 35-50 and > 51 years were 3.809 [AOR: 3.809, 95%CI: 1.61 - 9.02], and 3.59 [AOR: 3.59, 95%CI: 1.23 - 10.47] times higher as compared to household heads aged <35 years. On the other hands, the likelihoods were about 8.35 times higher for scheme enrolments for those HHs who walk ≤30 minute distances away to reach the nearest CBHI scheme contractual health care facilities from their residences [AOR: 8.35, 95%CI: 3.22 - 21.65] as compared to their counterparts in study area. Likewise, having positive attitudes towards CBHI scheme and its benefit package increases the HHs enrolments by 4.34 times more likely [AOR: 4.34, 95%CI: 1.525 - 12.32] than those HHs with negative attitudes toward the scheme in study area.

**Table 10:** Determinants enrollment to community-based health insurance obtained using multivariable logistic regression analysis among HHs (n=490) Chora District, Southwest Ethiopia, 2021.

| Variables code | Category | Participants |  | COR (95% C.I) | AOR (95% C.I) |
| --- | --- | --- | --- | --- | --- |
|  |  | Case n (%) | Control n (%) |  |  |
| Age (n=245) | <35 years | 66(26.9) | 142(58.0) | **1** | **1** |
|  | 35-50years | 118(48.2) | 69(28.2) | 3.67(2.43, 5.58) | 3.809(1.61, 9.02)<sup>a</sup> |
|  | >50 years | 61(24.9) | 34(13.9) | 3.86(2.32, 6.44) | 3.59(1.23, 10.47)<sup>a</sup> |
| Sex (n=245) | Male | 194 (79.2) | 226(92.2) | **1** | **1** |
|  | Female | 51 (20.8) | 19 (7.8) | 3.13(1.78, 5.47) | 10.85(3.01, 39.16)<sup>a</sup> |
| Family size (n=245) | $\leq 3$ | 15(6.1) | 95(38.8) | **1** | **1** |
|  | 4 – 5 | 79(30.2) | 101(41.2) | 5.59(3.02, 10.39) | 7.3(2.08, 25.84)<sup>a</sup> |
| | $\geq 6$ | 151(61.6) | 49(20.0) | 20.75(11.04, 38.9) | 13.31 (3.86, 45.92)<sup>a</sup> |
| Distance to health facility (n=245) | $\leq 30$ min' | 97(39.6) | 38(15.5) | 3.57(2.32,5.49) | 8.35(3.22, 21.65)<sup>a</sup> |
|  | >30 Min' | 148(60.4) | 207(84.5) | **1** | **1** |
| Perceived technical competences (n=245) | Favorable | 96 (39.2) | 45 (18.4) | 9.86(5.17,18.83) | 7.62(2.2, 26.4)<sup>a</sup> |
|  | Moderate | 133(54.3) | 126(51.4) | 4.88(2.699,8.83) | 2.46(0.8, 7.57) |
|  | Un-favorable | 16 (6.5) | 74(30.2) | **1** | **1** |
| Trust to CBHI Scheme management | Trusted | 84 (34.3) | 25 (10.2) | 12.82(6.93, 23.72) | 10.35(2.37, 45.17)<sup>b</sup> |
|  | Medium | 134(54.7) | 117(47.8) | 4.36(2.67, 7.14) | 4.91(1.9, 12.65)<sup>b</sup> |
| (n=245) | Not trusted | 27 (11.0) | 103(42.0) | 1 | 1 |
| Attitude towards<br>CBHI(n=245) | Positive | 176(71.8) | 42 (17.2) | 13.8(8.28, 22.99) | 4.34(1.53, 12.32)<sup>a</sup> |
|  | Neutral | 35 (14.3) | 91 (37.1) | 1.26(.733,2.18) | 0.36(0.12, 1.12) |
|  | Negative | 34 (13.9) | 112(45.7) | 1 | 1 |

<sup>b</sup>significant at 0.05, <sup>a</sup>significant at 0.01

## Discussion

The current study was aimed to identify factors that determine enrollment to community-based health insurance among households in Chora district, Southwest Ethiopia. Hence, the current study shows that being female-headed HHs increases the HHs scheme memberships by nearly 11 times more likely than male-headed households. This finding was consistent with studies conducted in North Gondar, Ethiopia, Ghana and Bangladesh (19,29,30). On the contrary, studies conducted in Northwest Ethiopia and Burkina Faso found that gender the household head has no association with the utilization of the CBHI scheme (28,31). The difference might be due to variation in population distribution from region to region and socio-demographic. Enrolments into the CBHI scheme were also determined by the age of the household heads; households who were above 35 years were more likely to be enrolled into CBHI scheme compared to adults below 35 years. This finding is supported with study conducted at North Gondar of Ethiopia, Southern Ethiopia and Tanzania (14,27,29). This association could be explained by the fact that household headed by older individuals could have larger family sizes including children and elders who need frequent health care services. Also, they might have more experience in visiting those health care facilities due to different medical conditions.. Consequently, they could have faced frequent financial hardship that might be happened due to direct OOPPs expenses that supported them to decide to be enrolled into CBHI scheme. However, a population-based case-control study conducted in rural Burkina Faso has reported young adults were more likely to be enrolled into CBHI scheme. (31).

Having large families were also found a predictor for CBHI enrolments in study area. Hence, as the family sizes increases the odds of enrolments increases. Therefore, this finding was supported with study conducted in Debub Bench, Sothern Ethiopia (27). Also, it was in line with the studies conducted in Bangladesh, India and Nigeria (14,30,32). The fact that household’s heads with larger family sizes who have elders and children are more likely to visit health facilities due to different illness of their loved once.. Nevertheless, this findings is contradicted with the study conducted in Burkina Faso and Bangladesh (33,34). This difference could be attributed to mechanisms of and amount of annual contribution by household heads. In Burkina Faso, the insurance scheme membership contribution was estimated individual per household. This had levied large amount of money as family size increased, and such kinds of families was battered, contrary to what has happened in Ethiopia. Until recent time, in Ethiopia, annual premium contribution is based at household level not individual level irrespective of family size (also called flat-rate payment system) (16,35–37).

Accessing a contractual health facility within a 30 minutes walking distance away from HHs’ residences increased the likelihood of enrollments in into CBHI scheme. This finding was supported with study findings from Debub Bench of Ethiopia; and mirrored with study conducted in rural Burkina Faso, Armenia, Benin, Ghana, and Rwanda(19,20,27,31,38,39). This effect might due to the fact that families who live at nearest distances to health care facilities are more likely to access health facilities and utilize health care services whenever they need.

On the other hand, the household heads who had positive perception toward public health care facilities health care service provider’s technical competencies ((40), and trustworthiness towards the scheme. (1,28,41–43) were more likely to enrolled into the CBHIs scheme as also reported by other study. The presence of such association might be due to the fact that household heads that had trusted the scheme might have previous good experience and trust in local organizations or other collective arrangements that could lead communities to trust with CBHI scheme management.

The current study also revealed that household heads that had positive attitudes towards the CBHI scheme were more likely to be enrolled into CBHI. This study was consistent with the study conducted in North Gondar, and Northeast in Ethiopia, Tanzania and Ghana (6,24,29,44).

Our study has several limitations it might be prone to information bias (specifically recall bias). Cases might remember more likely some exposure status (e.g. acute illness experiences) than controls. However, we have tried our best effort to minimize by shortening recalling time and using probing to remember past exposure experiences.

## Conclusion

This study demonstrated that the individual household heads related factors such as the individual household are age, who headed the household, and number of family sizes per households and attitudes towards the scheme were determinant risk factors of enrollment into CBHI scheme. From healthcare facility related factors: distance to arrive/reach at contractual health facility and perceptions towards the public health care providers technical competences, trust towards scheme managements, were determinant risk factors of enrollment into CBHI scheme. Therefore, we recommend household heads headed by females, has large family sizes should be given attention. Besides, it’s advisable to expand CBHI services to other health facilities for better access, improve service quality and scheme management system to increase trustworthiness of the users towards the CBHI scheme

## Data Availability

All the data produced in the present study are available upon reasonable request to the authors.

## Author Contributions

All authors contributed to data analysis, drafting or revising the article, gave final approval of the version to be published, agreed to the submitted journal, and agree to be accountable for all aspects of the work.

## Data Sharing Statement

The data were available from the corresponding author upon request.

## Funding

No specific fund was obtained to conduct this study.

## Disclosure

Authors declare that they have no competing interests.

## Acknowledgments

We would like to thank Jimma University for providing us the ethical clearance. We acknowledged the data collectors and study participants for their participation.

## Notes

### Competing Interest Statement

The authors have declared no competing interest.

### Funding Statement

This study did not receive any funding

### Author Declarations

Ethical approval was received from the institutional ethical review board of Jimma University, Institute of Health science.

